# Attention bias at baseline does not moderate the effect of attention bias modification for depressive symptoms

**DOI:** 10.1101/2024.11.10.24317072

**Authors:** Hallvard Solbø Hagen, Jan Ivar Røssberg, Catherine J. Harmer, Rune Jonassen, Nils Inge Landrø, Ragnhild Bø

**Affiliations:** Institute of Clinical Medicine, University of Oslo, Norway; Clinical Neuroscience Research Group, Department of Psychology, University of Oslo, Norway; Department of Psychiatry, Oxford University, UK; Faculty of Health Sciences, Oslo Metropolitan University, Norway

## Abstract

**Background:** Clinical trials of Attention Bias Modification for depressive symptoms have consistently produced small effect sizes and mixed results. Therefore, identifying subpopulations for whom this intervention works has been called for. Considering the intended mechanism behind Attention Bias Modification, change of attentional bias, the level of bias at baseline may moderate its efficacy.

**Methods:** Participants with a history of depression (N=301) were randomized to receive two daily sessions of either Attention Bias Modification or sham for 14 days. A response-based attention bias score was calculated, and a moderator analysis was run at post-intervention and 1-month follow-up measured by change in Hamilton Depression Rating Scale and Beck Depression Inventory II, respectively.

**Results:** Baseline attention bias did not significantly moderate the effect of Attention Bias Modification on any of the time points or depression measures.

**Conclusions:** Baseline attentional bias was not found useful for characterizing subgroups more likely to benefit from ABM for depressive symptoms.

## Introduction

Depression is a leading cause of disability worldwide (GBD 2019 Mental Disorders Collaborators 2022). Current treatment options are effective [2], but a considerable proportion of patients experience residual symptoms and frequent relapses. Therefore, to improve treatment outcomes, novel approaches are required. Attention Bias (AB), the selective attention to negatively valenced information, is theorized to be a central mechanism of depression [3]. Meta-analyses show that patients with both current (Peckham et al., 2010; Suslow et al., 2020) and remitted depression [6,7] show AB for negative stimuli.

Attention Bias Modification (ABM) was developed to reduce negative AB. In a seminal study, it was revealed that modification of AB decreased mood reactivity following a mood induction procedure [8]. In the following decades, ABM has been found to decrease depressive symptoms (e.g. Hsu et al., 2021; Jonassen et al., 2019) but effects have been small or non-significant in the early meta-analyses [11–13]. Two more recent depression-specific meta-analyses showed small to moderate effect sizes [14,15].

Under the requirements of precision medicine, a more fine-grained approach for personalizing treatment has been advocated by the Lancet Commission of Depression [16]. Various potential moderators have been put to the test to define participant characteristics that may predict the treatment outcome of ABM. In anxiety research, several moderators have been identified (MacLeod et al., 2019), but few moderators have been found when investigating depression. A higher baseline score of anxiety [18] and baseline rumination [19] have been found to moderate the effect, though non-significant moderator effects have also been reported [4]. A major problem in some of these studies has been lack of statistical power.

Change in AB is the assumed causative agent of ABM and we therefore sought to analyze whether the baseline level of AB would moderate the effect of ABM. A significant moderating effect of baseline AB has been found in generalized social phobia [20], but has not, to our knowledge, been tested appropriately in depressed patients. In a study on non-depressed college students, a moderating effect was absent in a three-armed RCT (N=77), using a dot-probe task [21]. However, pre-selecting individuals based on moderate levels of AB in eye-tracking in a group of depressed individuals was found to have a lasting effect on depression [9], but moderation was not analyzed. This may indicate a more beneficial effect for individuals with more negative ABs.

The reliability of the traditional AB computation has repeatedly been found to be unacceptably low [22–25]. This makes it both unapt for clinical use as it cannot be used at an individual level but also reduces power [26]. We therefore applied a novel computational model that has been shown to increase reliability to acceptable levels [27]. Rather than a total trial average of reaction times, this response-based computation is more sensitive to fluctuations in attention. From our material, three of the indexes can be calculated: vigilance, avoidance and the ratio between the absolute numbers. As the group mean AB has been found to be toward dysphoric images across different measurement techniques [5,7], we chose vigilance as the metric of AB in this analysis.

Baseline AB could be used as an indicator of treatment strategy and this could help tailor treatment, and consequently a more personalized treatment. We hypothesized that the baseline level of AB in a sample of previously depressed participants would moderate the effect of ABM on depression outcome with increasing effect for those displaying more extreme baseline AB scores.

## Methods

### Participants

The data analyzed in this paper is based on a previous double-blinded randomized controlled trial including participants with residual depressive symptoms with preregistered moderator analyses. Participants were recruited after receiving treatment as outpatients and were included in the study between May 6, 2014 to September 27, 2016 (ClinicalTrials.gov, number NCT02658682; n=301) [10]. The study received approval from the Regional Ethical Committee for Medical and Health Research for Southern Norway (2014/217/REK Sør-Øst D) and informed written consent was obtained prior to enrollment. Participants aged 18-65 were included if they, according to the Norwegian version of MINI International Neuropsychiatric Interview 6.0.0 (M.I.N.I.), met the criteria for recurrent remitted Major Depressive Disorder (MDD), and having at least two prior episodes of depression. Exclusion criteria were head trauma, current substance abuse, psychosis, bipolar disorder, neurological disorders, and attention deficit disorders. The data was accessed in March 2024.

### Randomization

Participants were randomly allocated 1:1 to receive either active ABM or a sham condition and were provided with laptops to run the intervention at home. The laptops were pre-programmed by an independent lab technician, thus ensuring blindness to study allocation for both participants and outcome assessors. The study followed an ITT approach, implying that 37 participants who were currently depressed and therefore wrongfully included were subjected to analysis.

### Intervention

The ABM procedure was based on the dot probe procedure and was similar to the one described in Browning et al. (2012). Participants were instructed to perform the task, consisting of 96 trials and lasting for approx. 5-7 min, twice daily for 14 days. Each trial started with a fixation cross, followed by a vertically positioned pair of images of faces and these were paired positive-negative, positive-neutral and negative-neutral. All pictures were from 4 databases: the Karolinska directed emotional faces database (Lundqvist et al., 1998), Matsumoto and Ekman’s Japanese and Caucasian Facial Expressions of Emotion (Biehl et al., 1997) NimStim (Tottenham et al., 2009) and Ekman Pictures of Facial Affect (Ekman & Friesen, 1976). The stimuli lasted 500ms or 1000ms (1:1) and was randomly presented in either the first or last half of the session. The emotions expressed were fear, anger, happiness, and neutrality. The pictures were immediately followed by a single or pair of dots (the probe) which replaced the more positively valenced facial expression 87% of the time. The participants were instructed to identify the number of dots as quickly and accurately as possible by pressing one of two keys on the keyboard. The sham condition was identical except for a 50% chance of the dot replacing either valence.

### Assessments and outcomes

Hamilton Depression Rating Scale (HDRS; Hamilton, 1960) and Beck Depression Inventory-II (BDI-II; Beck et al., 1988) were used as outcomes and these were measured at baseline, immediately after the two-week intervention, and 1 month post-intervention. These showed good internal consistency with Cronbach’s Alpha at 0.771, 0.813 and 0.814 (HDRS) and 0.918, 0.927 and 0.931 (BDI-II) respectively. Reviewers underwent training using case examples. Throughout the trial, bi-weekly supervision meetings were held to maintain consistent rating standards. Any deviations were resolved through consensus among the reviewers. However, we did not formally assess the ICC between interviewers.

Baseline AB was measured in a paradigm equal to a single round of the sham condition but with a new set of facial stimuli. Reaction times (RT) for responding to the probe by pressing one of two designated keys on the keyboard were measured. The more negative expressions (e.g. neutral vs positive) was defined as stimulus, thus defining valid trials as the ones were the dot-probe replaced the more negative expression. In the invalid trials, the dot-probe replaced the least negative picture. As there were no neutral-neutral trials, only the vigilance and avoidance scores were calculated. Each individual reaction time of the valid scores was subtracted from the averaged score of the invalid trials. Positive difference scores were averaged per participant and termed “vigilance” and negative difference scores were averaged per participant and termed “avoidance”. A vigilance-avoidance ratio was also calculated. For comparison, traditional AB scores were also included where the mean RT of the valid negative trials was subtracted from the mean RT of the invalid scores.

### Statistical analysis

Statistical analyses were performed using R[31] in RStudio v2023.12.0[32]. There were missing data sets on HDRS 1-month (n=30 (10%)) and BDI-II 1-month (n=28 (9%)). These were excluded from the 1 month analyses. Missing data points (n=13) were replaced with a series mean as they were missing completely at random. Descriptives, including correlation analyses, were conducted for baseline measurements. A regression-based moderator analysis was performed using the PROCESS macro v4.3 [33] and assumptions were tested and met according to Hayes [34]. The predictor was set as ABM, the moderator as baseline AB and outcome as BDI-II and HDRS at post-intervention or 1 month post-intervention. The threshold for significance was set at 0.05. If significant, the interaction terms were probed at the 16^th^, 50^th^ and 84^th^ percentiles and regions of significance were examined using the Johnson-Neyman technique. Bootstrapping was performed to obtain robust confidence intervals for the interaction term (n=5000). All regression analyses were controlled for the respective depression score at baseline.

The attention bias data was reduced by first removing trials outside 200-2000ms. Next, answers with the wrong location were removed. Missing data was imputed using series mean. The reduction left 96.23% of the trials. No data points were considered multivariate outliers according to the criteria of being positive on 2 out 3 on Mahalanobi’s distance, Cook’s distance and/or centered leverage, as applied in [18]. After randomization, 37 individuals were found to fill the criteria for MDD (MDD+), thus violating the inclusion criteria. Explorative analyses were run for the MDD+ group and selecting for antidepressant status.

Reliability for the response-based score was measured using split-half analysis using even and odd trial numbers to account for training effects [35]. All reliability correlation coefficients were calculated using Spearman’s rho and corrected with the Spearman-Brown prophecy formula. A sensitivity power analysis was calculated using GPower [36].

## Results

### Baseline characteristics

Demographics are shown in Table 1. A correlation matrix between study variables is shown in Table 2. HDRS and BDI showed a significant correlation coefficient of 0.65, but response-based AB at baseline showed a non-significant correlation of 0.05 with BDI-II and 0.02 with HDRS. Mean negative vigilance was 88.7ms (31.1ms), mean negative avoidance -131ms (57.9ms) and the vigilance-avoidance-ratio was 0.768 (0.418). Analyzing baseline AB by status of depression with independent two-sided t-test showed a significantly higher value for vigilance in the Major Depressive Disorder subgroup (MDD+) (MDD+: 100.0 ms vs MDD-: 87.1 ms, p=0.03) but non-significant differences in mean avoidance (MDD+ -138.0 ms (54.9ms) vs MDD-130.0 ms (58.3ms), p=0.422). Eighty participants were using SSRIs or SNRIs (AD+), but there was no difference in mean vigilance (AD+: 91.9ms(28.9ms) vs AD-:87.5(31.9ms), p=0.30) or in mean avoidance in these groups (AD+: - 133ms(58.3ms) vs AD-:-130ms(57.4ms), p= 0.65). Traditional score AB at baseline across the sample was not significantly different from zero (mean -1.07ms, CI [-4.25 – 2.12], t(301)=-0.659, p=0.51).

**Table 1.** Demographics (means (SD) or frequencies).

|  | Placebo (n=148) | ABM (n=153) |
| --- | --- | --- |
| Sex (females) | 103 | 109 |
| Age | 41.5 (13.6) | 40.2 (12.7) |
| Education level (ISCED) <sup>a</sup> | 6.0 (1.2) | 6.0 (1.1) |
| HDRS pre-intervention | 8.3 (5.1) | 9.3 (6,0) |
| HDRS post-intervention | 8.7 (5.7) | 8.4 (6,0) |
| HDRS post 1 month | 8.1 (5.7) | 8.6 (5.9) |
| BDI-II pre-intervention | 13.5 (9.7) | 15.1 (10.6) |
| BDI-II post-intervention | 11.0 (9.7) | 11.5 (10.5) |
| BDI-II post 1 month <sup>b</sup> | 10.6 (9.5) | 12.3 (10.1) |
| AB bl traditional ms | -1.2 (28.0) | -0.9 (28.0) |
| AB bl vigilance ms | 87.0 (33.7) | 90.1 (28.2) |
| AB bl avoidance ms | -130.0 (55.0) | -131.0 (60.3) |
| AB vigilance-avoidance ratio | 0.7 (0.1) | 0.7 (0.2) |
| Compliance<sup>c</sup> | 80.0% (21.5) | 83.4% (15.0) |

Note. AB = Attentional Bias. ABM = Attention Bias Modification. BDI-II = Beck Depression Inventory II. HDRS =Hamilton Depression Rating Scale. bl = baseline.
<sup>a</sup> Missing data for 6 in the placebo group, 8 in the ABM group
<sup>b</sup> Missing data for 17 in the placebo group and 11 in the ABM group
<sup>c</sup> percentage of max 2688 trials

**Table 2.**
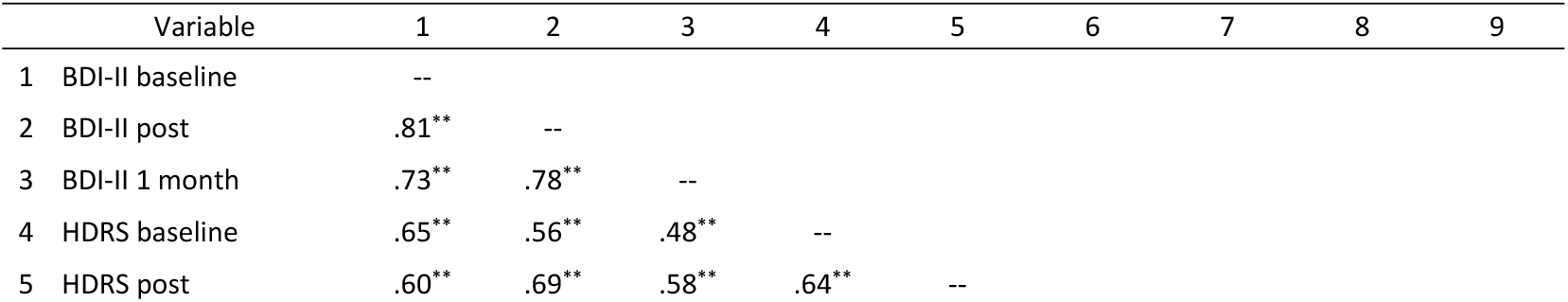
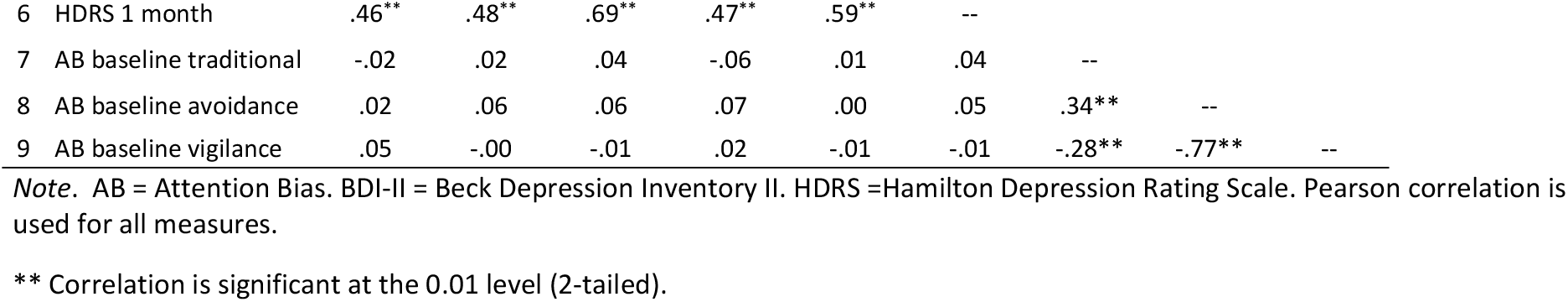
Correlation matrix of psychometric scores.

The dot-probe reaction times showed high split-half reliability (valid: r=0.96, invalid: r = 0.97) and the correlation between mean average RT for valid and invalid trials were high (r=0.96). The split-half reliability for average vigilance was 0.86 and 0.54 for avoidance, respectively. Subanalyses for stimulus duration showed a reduction in reliability for both durations in vigilance (500ms: mean r=0.70, 1000ms: mean r=0.74) and increase at 1000ms in avoidance (500ms: mean r: 0.48, 1000ms: mean r: 0.58). Based on the reliability measurement, vigilance was used as measure of AB in the moderator analyses.

### Moderation analysis

There was a statistically significant improvement in HDRS at post-intervention in the intervention group compared to the sham group (F (1,309) = 6.78, η2 = .02, p < .01], see Jonassen et al 2019 for further details). Response-based baseline vigilance AB scores did not moderate the effect of ABM on change in any of the outcomes.

For HDRS at post-intervention: F(4,295)=13.702, p<0.001, MSE=20.242. R^2^-change: 0.001, F(1,295)=0.303, p=0.582, controlling for baseline HDRS. The slopes of the interaction effect are represented in Fig 1. The bootstrapping confidence interval for HDRS post-intervention was - 0.024 - 0.037, i.e., covering zero.

**Fig 1.**
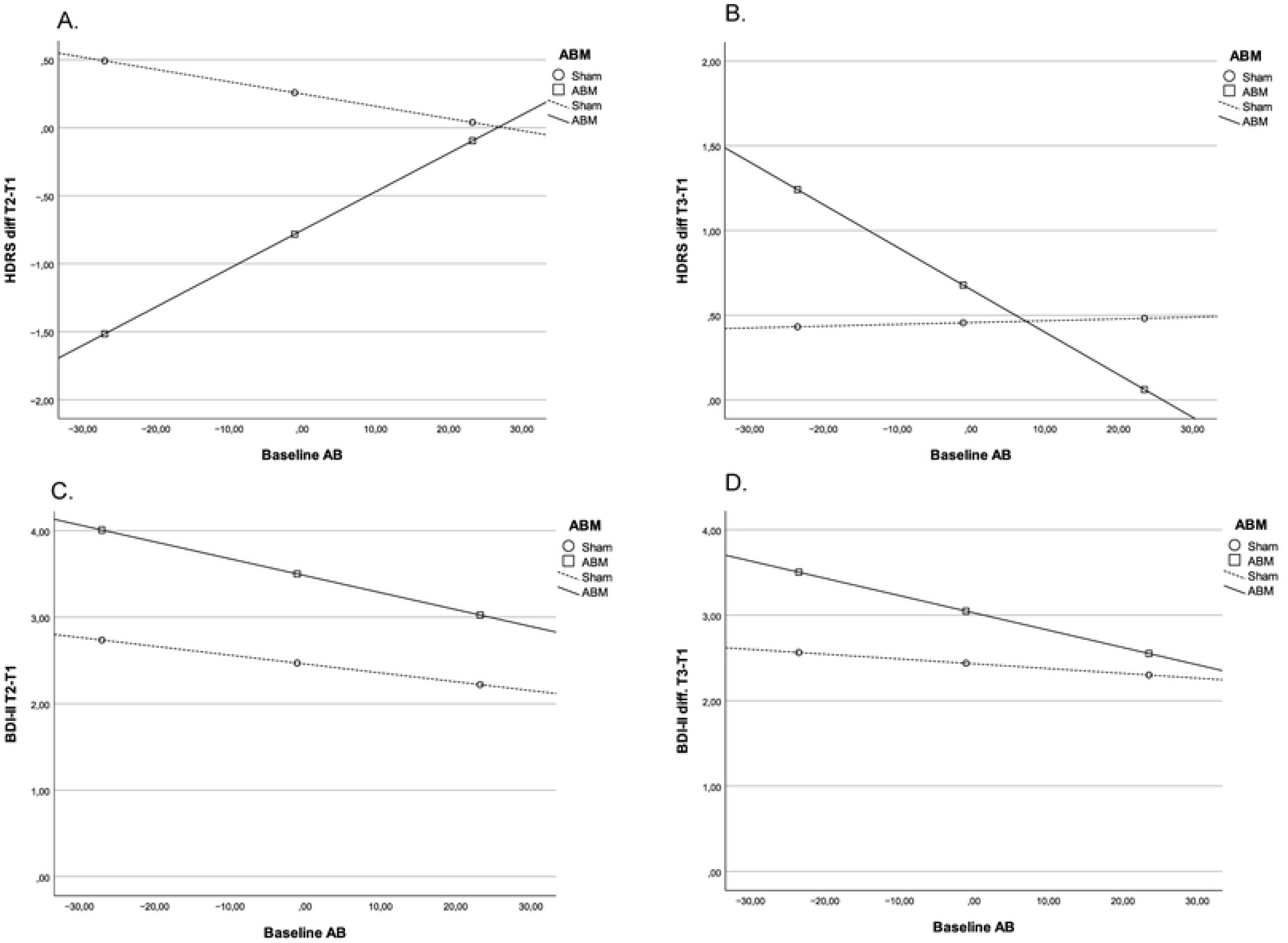
Moderator analysis of baseline AB on depression outcomes. *Note*. **A**. Change in HDRS from baseline to post-intervention. **B**. Change in HDRS from baseline to 1 month. **C**. Change in BDI-II from baseline to post-intervention. **D**. Change in BDI-II from baseline to 1 month. Neither of the interaction effects were significant. ABM = Attention Bias Modification. BDI-II = Beck Depression Inventory II. HDRS =Hamilton Depression Rating Scale. RRS-B Rumination Response Scale – Brooding subscale

Baseline vigilance AB scores did not moderate the effect of ABM on reduction in BDI at post-intervention: F(4,295)=0.949, p<0.0001, MSE:36.107. R^2^-change: 0.004, F(1,295)=1.1567, p=0.283.

Bootstrap CI: -0.014-0.0669. Excluding the (MDD+) did not affect the results.

Baseline vigilance AB scores did not moderate the effect of ABM on change in HDRS at 1 month follow-up: F(4, 265)=22.719, p<0.001, MSE: 26.242. R^2^-change 0.000, F(1,265)=0.000, p=0.247.

Bootstrap CI: -0.070-0.016. Excluding the (MDD+) did not affect the results.

Baseline AB did not moderate the effect of ABM on change in BDI at 1 month follow-up: F(4,267)=13.361, p<0.001, MSE:44.700. R^2^-change: 0.001, F(1,267)=0.021, p=0.885.

Bootstrap CI: -0.050-0.041. Excluding the (MDD+) did not affect the result. For full results, see supplementary information.

## Discussion

The present study investigated response-based baseline AB as moderator for the effect of ABM on depressive symptoms in a sample of participants with residual symptoms. No moderator effect was found in either depression outcome at post-intervention or after 1 month.

Given the assertion that the central mechanism of ABM is correction of AB, the lack of a moderating effect of baseline AB is surprising [37]. Still, this is in line with the only other moderator study on baseline AB, where a negative AB at baseline did not moderate the change in depression in a non-clinical population [21]. These results might represent a false negative as the analysis of moderation probably was numerically underpowered (three arms, n=77) in addition to the use of the unreliable traditional AB score. On the other hand, pre-selecting for a group with a moderate negative baseline using a more reliable score, Hsu et al (2021) gave a significant effect of 0.72 points reduction per week over four weeks on the HRSD. This could indicate an effect of pre-selecting on AB, but as no control group was included, this remains uncertain.

Baseline AB as moderator for the effect of ABM has been more extensively studied in anxiety, though results are mixed. Amir et al (2011) used a dot-probe task and found baseline AB towards threat to moderate the response to the intervention in individuals with generalized social phobia. On the other hand, in a study including children with social anxiety disorder, no moderating effect of baseline AB was found on clinical outcome [38].

There are several explanations for the lack of moderation effect in our study. First, there was no significant mean negative traditional AB at baseline. This is surprising, considering the results of a meta-analysis where previously depressed individuals were found to have a negative traditional AB, comparable to the AB found among currently depressed individuals [7]. Still, in never-depressed people a neutral average AB is not the norm, but rather a positive bias [39]. As such, the level in our sample represents a negative deviation from never-depressed.

Second, though the response-based AB calculation showed high reliability, it was not correlated with any of the baseline depression scores, thus possibly being of questionable validity as a measurement of depression. This corroborates previous findings on depression scores and AB measurement [40,41]. The group differences in AB between depressed and non-depressed has been a consistent finding and thus leaves the question of what the relationship between AB and depression represents. The use of sum-scores as outcome may be conceal the effect as depression represents a highly heterogenous diagnosis but no subscale to link depression to attention bias has been identified or agreed [40,42].

Third, some choices in the baseline AB measurement paradigm may have affected the result. In Kaiser et al (2018), the combination of emotionally congruent information in combination with self-referential cues were the most potent at eliciting negative AB. The importance of relevance to the participant was found for positive biases in Pool et al (2016). The dot-probe task may be less apt at assessing AB due to the lack of self-referential material in our study. Stimulus lengths have also been found to affect the AB detection. In our study, a stimulus length of 500 ms and 1000 ms was set at a ratio of 1:1. Donaldson et al (2007) found a significant negative baseline AB in depressed individuals when extending stimulus presentations to 1000 ms, which was not found at 500 ms. Further, eye-tracking-studies have supported the notion of a bias in the latter stages of attention in depression [5]. If AB is dynamic, rather than static, mean reaction times derived from longer stimulus durations may provide a more reliable measurement, such as those found in eye-tracking studies (Skinner et al., 2018).

Fourth, the low depression levels in the current sample may be of relevance to the null finding. With a baseline BDI-II score of 14.3 in the sample, smaller improvements would be expected than in a clinically depressed sample. Still, a baseline AB has been found in samples with lower baseline BDI scores. In Arenliu et al (2023), the baseline score of BDI-II in a non-clinical population was 11.4, lower than our baseline, and still a clear AB was found, though by using eye tracking instead of dot-probe. In another sample, Joorman and Gotlib (2007) found a baseline AB in both depressed and formerly depressed patients, the latter with a mean BDI of 5.21, using a dot-probe stimulation period of 1000ms. It is therefore unclear whether the low level of depression may have affected the result, but limited opportunity for change in the outcome will anyhow limit the variance to explain.

Fifth, 27% (n=80) of the current sample used antidepressant medication, which could partly explain the small negative traditional AB. SSRIs have been found to reduce attentional vigilance to threat in healthy volunteers [46] but has to our knowledge not been studied in a clinically depressed population Likewise, differences in AB using eye-tracking also revealed differences between medicated and non-medicated individuals with MDD [47]. Still, considering the non-significant difference in baseline response-based AB between antidepressant-users and non-users, the effect does not seem to be the explain the lack of negative AB.

It is difficult to know if previous psychotherapy affected the baseline measurement here. The research is scarce but a significant reduction after both CBT and positive psychology interventions in AB using eye-tracking technology has been found [48]. Being a different detection method, it is not possible to translate these findings to our study as the detection methods have been found to be largely uncorrelated [25]. Furthermore, information of the types of pre-intervention psychotherapy was not collected.

Sixth, the lack of moderation effect may be attributed to the limited effect of the intervention and the similarity of the sham comparator. The total change in HDRS for the intervention group was small (−0.9 points vs +0.5 for the sham condition) and not significant for BDI-II, possibly due to the low baseline depression levels. The use of a sham condition as control to address the clinical utility of ABM has been questioned because it is not a clinically relevant alternative [49] and also due to the effect on attentional processes by both conditions [22,50]. It may also alter moderating factors of AB such as attentional control (Basanovic and MacLeod, 2017) and improvement in attentional control, following ABM, has been found to correlate with clinical improvement [22,50]. It may therefore mask the effect by itself affecting AB, reducing the chance of finding a statistically significant result. Still, Hsu et al (2021), pre-selecting for moderate AB, found a significant difference in depression scores between sham and active ABM, but no difference between sham and assessment-only.

Seventh, in this study there was a significantly larger change in HDRS in the ABM group than the control group, but the change in the traditional AB was small and not significantly, as previously reported [10]. Macleod and Grafton (2016) showed how a lack of significant change in AB often resulted in lack of change in depression outcomes. The eliciting of a significant change in AB is necessary to determine if the procedure was effective, rather than making claims about the effect of the process [52]. This may have affected the size of the reduction and may be a consequence of the mentioned similarity in conditions.

### Strengths and limitations

This RCT represents the largest RCT in individuals with residual depression, using a well-known procedure with stringent conditions. In addressing the residual nature of depression, it answers an important question of whether using baseline AB moderates the effect of ABM. By applying the response-based AB measure, a high reliability secures the frequently overlooked problem of poor reliability on power calculations [26], and the consequent underestimation of the number of participants needed. As this is secondary analysis no a priori power calculation was performed, but a sensitivity power analysis [53] using GPower 3.1.9.7 fixing α= 0.05 and power at 0.90 with n=301 returns a possible detection of effect sizes of f^2^= 0.04 indicating an unlikely false negative result [36].

Generalization to other populations is limited due to lack of generalizability due to low baseline depression scores. Another issue lies in the lack of sad facial expressions in the stimuli as this has shown to be the most potent stimuli in eliciting AB in depression. Furthermore, the analysis of single moderators is possibly not sufficient to define the treatment of MDD [54] and multivariate approaches may be more clinically useful, such as the Personalized Advantage Index that has shown promising results in psychotherapy research [55].

### Future directions

As previously suggested, the validity of the response-based index should be studied and outcome measures evaluated further in clinically depressed populations [40]. Further, we support the notion that both cleaning of data and reliability measure should be registered before inclusion as suggested by Molloy & Anderson (2020). Furthermore, the use of a sham condition as control group in clinical trials should be carefully considered due to its similarity to the active intervention.

## Conclusion

A response-based baseline measure of attention bias did not moderate the effect of ABM despite a reliable AB measure, thus rendering it unsuited to define individuals with increased effect of ABM.

Furthermore, it was not correlated with levels of depression and further research to clarify this relationship and the psychometric validity is needed.

## Data Availability

The dataset analyzed during the current study is not publicly available due to privacy reasons but is available from the corresponding author on reasonable request and with necessary ethical approval.

## Acknowledgements

**Acknowledgements:** We want to thank Luigi Maglanoc, Eva Hilland, Inger Marie Andreassen, Adrian Dahl Askelund, Dani Beck, Sandra Aakjær Bruun, Jenny Tveit Kojan, Nils Eivind Holth Landrø, Elise Solbu Kleven and Julie Wasmuth for their contribution to the data collection. Further, we thank Kari Agnes Myhre, Erlend Bøen, MD, PhD and Torkil Berge at Diakonhjemmet Hospital, Division of Psychiatry, for help and support during the recruiting period. We also thank our external recruitment sites, Unicare, Tore C. Stiles at Coperiosenteret AS, Torgny Syrstad, MD, Synergi Helse AS and Lovisenberg Hospital.

